# High-Performance Classification of Mpox Symptoms Using Support Vector Classifier and Quadratic Discriminant Analysis

**DOI:** 10.64898/2026.02.12.26346046

**Authors:** Shalom. C. Okoli, Funmilayo. C. Ligali, Michael Olufemi, Kolapo Oyebola

## Abstract

**Background:** Recent global outbreaks of Mpox have posed significant diagnostic challenges, particularly in resource-limited settings. Conventional diagnostic methods are often inaccessible due to cost, logistical constraints, or lack of trained personnel. These limitations highlight the urgent need for alternative, scalable diagnostic strategies. This study explored the application of machine learning (ML) classifiers trained on clinical symptom data as a rapid, cost-effective tool for Mpox detection.

**Methods:** An open-access dataset of clinical symptoms from suspected Mpox cases was used to train and evaluate five supervised ML algorithms: Extra Trees, Quadratic Discriminant Analysis (QDA), Decision Trees, Perceptron, and Support Vector Classifier (SVC). Prior to training, data preprocessing steps, including normalization and handling of missing values, were performed after which model training was carried out using a stratified 80:20 train-test split. Performance was assessed using accuracy, recall, area under the receiver operating characteristic curve (ROC-AUC), and F1-score metrics. Subsequently, feature importance was analyzed using permutation-based techniques to determine the contribution of each clinical symptom to model predictions.

**Results:** Among the five evaluated models, SVC, QDA, and Perceptron achieved superior and identical performance metrics, with accuracy, ROC-AUC, and F1-score values of 97.7%, and a recall of 95.5%. Each of these models correctly identified 44 true positive cases with zero false positives. In addition, QDA and SVC produced the lowest number of false negatives (2) and the highest number of true negatives (42), indicating robust discriminatory power. Feature importance analysis identified skin rash as the most predictive clinical feature, with a permutation importance score of 0.12.

**Conclusions:** These findings demonstrate the strong potential of machine learning classifiers for detecting Mpox based on clinical features. Incorporating these models into healthcare systems could significantly enhance early case detection, improve clinical decision-making, and bolster disease surveillance. Future research should focus on prospective validation of these ML classifiers in real-world clinical environments.

## Background

Monkeypox virus (MPXV) is a double-stranded DNA virus that belongs to the genus *Orthopoxvirus* within the family *Poxviridae*, and is the etiological agent of the disease monkeypox (Mpox) [1, 2]. Endemic to the rainforests of Central and West Africa, MPXV is classified into two genetically distinct clades: Clade I (Congo Basin) and Clade II (West Africa), with the latter further divided into subclades IIa and IIb [3, 4]. Clade I is associated with higher virulence and mortality rates, whereas Clade II is generally linked to milder disease outcomes. Notably, recent outbreaks, including the 2024 epidemic, have been primarily driven by Clade I subclade IB, which exhibits increased human-to-human transmission and elevated mortality [5, 6].

Mpox typically presents with fever, headache, lymphadenopathy, and a characteristic rash that evolves from macules to vesicles and pustules. These symptoms closely resemble other exanthematous illnesses such as measles, smallpox, and varicella (chickenpox), complicating differential diagnosis [7]. Other conditions commonly misidentified as Mpox include bacterial skin infections, scabies, and syphilis. In endemic regions, as many as 50% of suspected Mpox cases are misdiagnosed as chickenpox [8].

Laboratory confirmation is traditionally achieved through the detection of viral DNA from lesion specimens via conventional or real-time polymerase chain reaction (PCR), and sometimes confirmed through sequencing [4, 9]. However, these molecular diagnostic techniques are costly, labor-intensive, and largely inaccessible in remote or resource-constrained settings. For instance, during the 2023 Mpox outbreak in the Democratic Republic of Congo (DRC), only about 9% of suspected cases underwent PCR testing, with most diagnoses made based on clinical presentation alone [10].

Despite international recommendations calling for laboratory confirmation—especially following the global surge in cases after 2022—many endemic countries continue to rely on clinical diagnosis due to infrastructural and financial barriers. In 2023, out of approximately 14,000 suspected Mpox cases in the DRC, fewer than 2,000 were laboratory-confirmed [11]. Underdiagnosis and underreporting, often driven by misdiagnosis, limited diagnostic infrastructure, poor surveillance systems, and stigma, significantly hinder effective public health responses in Clade I-affected regions [12].

These limitations underscore the urgent need for accessible, cost-effective, and non-invasive diagnostic alternatives. Artificial Intelligence (AI)-based approaches, particularly machine learning (ML) models, have shown promise in epidemic surveillance and disease prediction [13, 14]. While earlier studies have largely focused on deep learning methods applied to skin lesion imagery [15, 16], few have explored the diagnostic potential of clinical symptom data alone. In this study, we investigated the performance of supervised machine learning classifiers for Mpox prediction using openly accessible symptomatology datasets.

## Methods

### Data Collection

All analyses were conducted in Google Colab using Python 3.11.11, following the analytical workflow outlined in Figure 1, which included stages of data acquisition, cleaning, preprocessing, feature engineering, exploratory analysis, and supervised machine learning classification. A line-listed dataset of suspected and confirmed Mpox cases reported from 2022 onward was obtained from the World Health Organization through the Global.Health data platform, accessed on 28 October 2024 [17]. The dataset provided anonymized individual-level case data.

**Figure 1.**
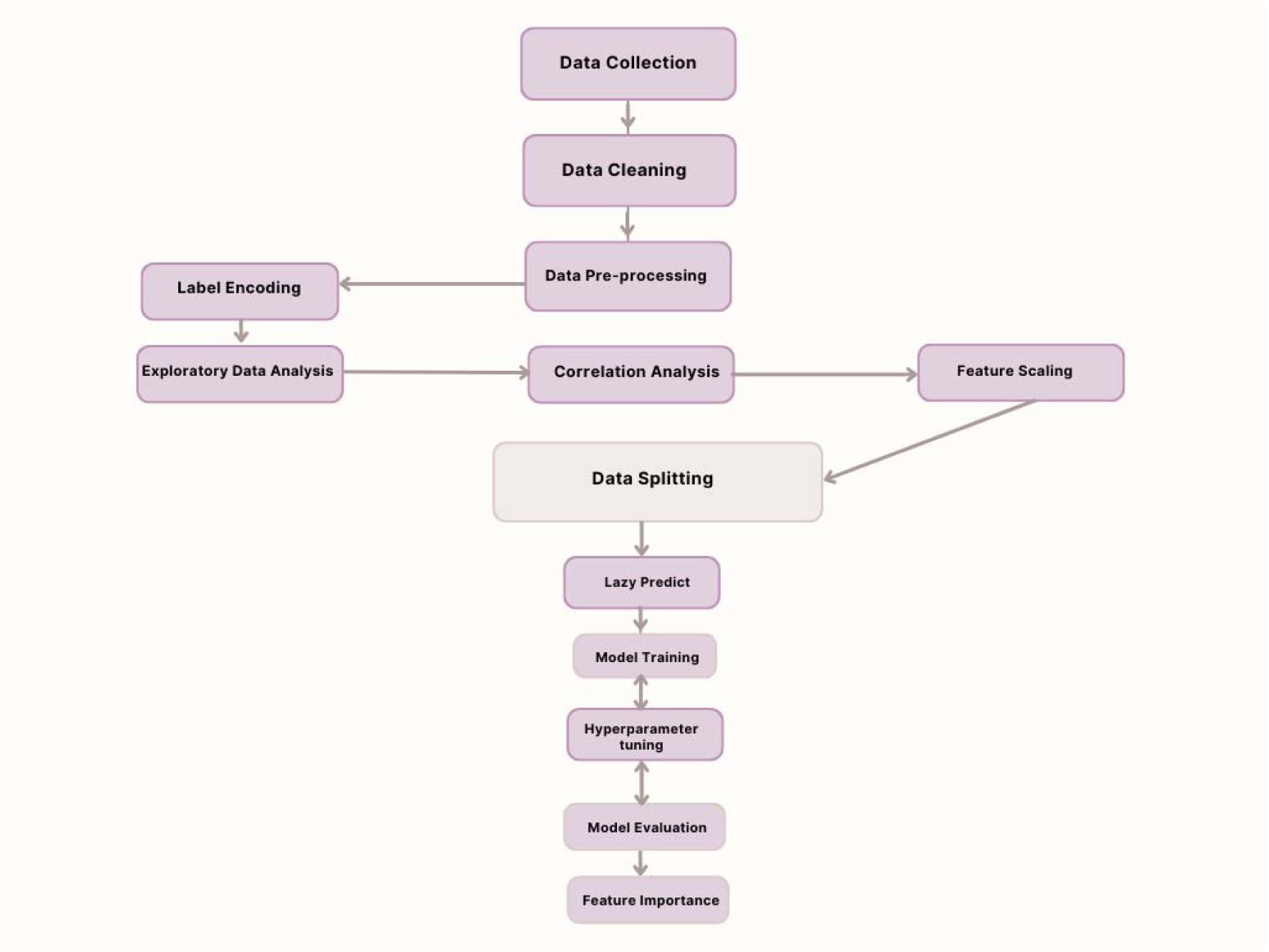
Workflow for supervised machine learning analysis.

### Data Cleaning

The original datasets (https://github.com/globaldothealth/outbreak-data/wiki/GHL2024.D11.1E71 and https://github.com/globaldothealth/monkeypox) comprised variables such as gender, age, disease status, geographic location, and reported symptoms, in addition to 22 other variables deemed extraneous to this analysis. Due to variability in data quality and missing values across countries, a refined dataset was curated, retaining only essential columns: ‘Status’, ‘Country’, ‘Age’, ‘Gender’, and all symptom-related entries.

Symptom variables were highly heterogeneous in terminology and format. To ensure consistency, similar symptom descriptors were standardized (e.g., ‘muscle pain’, ‘muscle ache’, and ‘myalgia’ were harmonized as ‘muscle ache’) [18]. Symptom values were binarized (i.e., ‘Yes’ = 1, ‘No’ = 0), and the ‘Status’ variable was encoded as Confirmed = 1 and Suspected = 0. A new column, ‘Transformed Age’, was created to group individuals into three age brackets: 0–19, 20–64, and 65 and above, in line with public health stratification practices [19].

To mitigate class imbalance, an equal number of suspected cases were randomly sampled to match the number of confirmed cases. The final balanced dataset consisted of 438 individuals with 48 columns: 42 symptoms, and six demographic or administrative variables including ID, Status, Country, Age, Transformed Age, and Gender (Table 1).

**Table 1:** Sample Dataset after Data Cleaning (due to its size, this table has been uploaded as an additional file)

### Data Preprocessing and Feature Engineering

Missing entries in the ‘Gender’ and ‘Age’ fields were imputed using the mode of the respective variables, a common practice for categorical and semi-continuous attributes [20]. Duplicate entries were identified and removed. For model compatibility, the ‘Status’ column was relocated to the final position in the dataset. Categorical features such as Country and Gender were label-encoded using Scikit-learn LabelEncoder module [21].

### Dataset Splitting and Feature Scaling

The dataset was partitioned into training, validation, and test subsets in an 80:10:10 ratio using the train_test_splitfunction from Scikit-learn. Prior to model fitting, all features were standardized using z-score normalization (mean = 0, standard deviation = 1) to ensure comparability across machine learning algorithms [22].

### Correlation Analysis

Correlation matrices were generated to explore linear relationships between symptoms and to evaluate associations between each feature and Mpox status. These were visualized using heatmaps (Figures 2A and 2B), offering insights into symptom clusters and potentially predictive features.

**Fig 2A:**
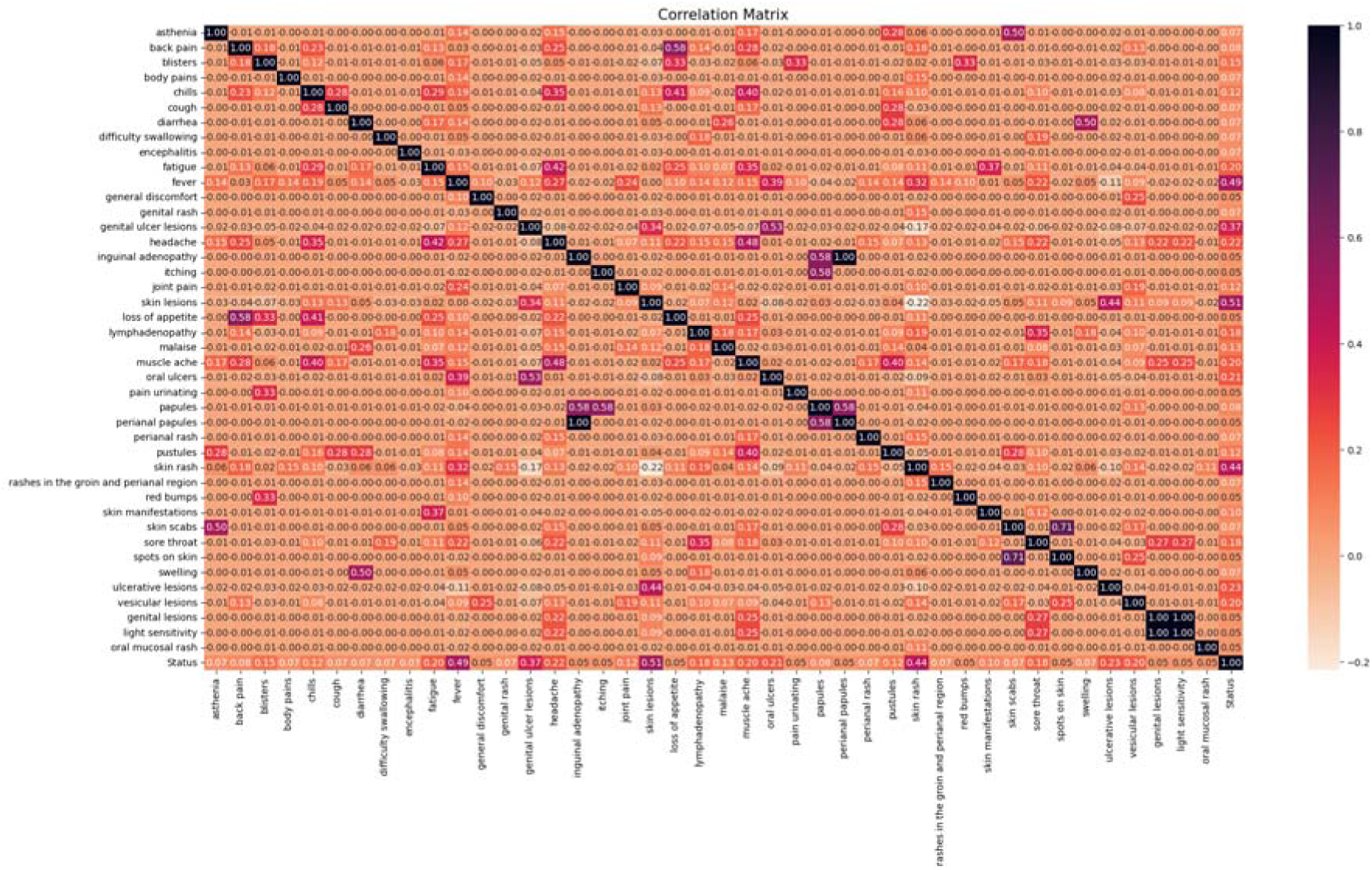
Correlation Matrix of features (symptoms) in the dataset. Values range from −1 to 1; where −1 indicates perfect negative correlation and 1 indicates perfect positive correlation. Diagonal black boxes across the matrix show direct correlation between features. Strong positive correlation is shown in purple graded boxes while strong negative correlations are shown in red graded boxes. Colour intensities are directly proportional to correlation scores.

**Fig 2B:**
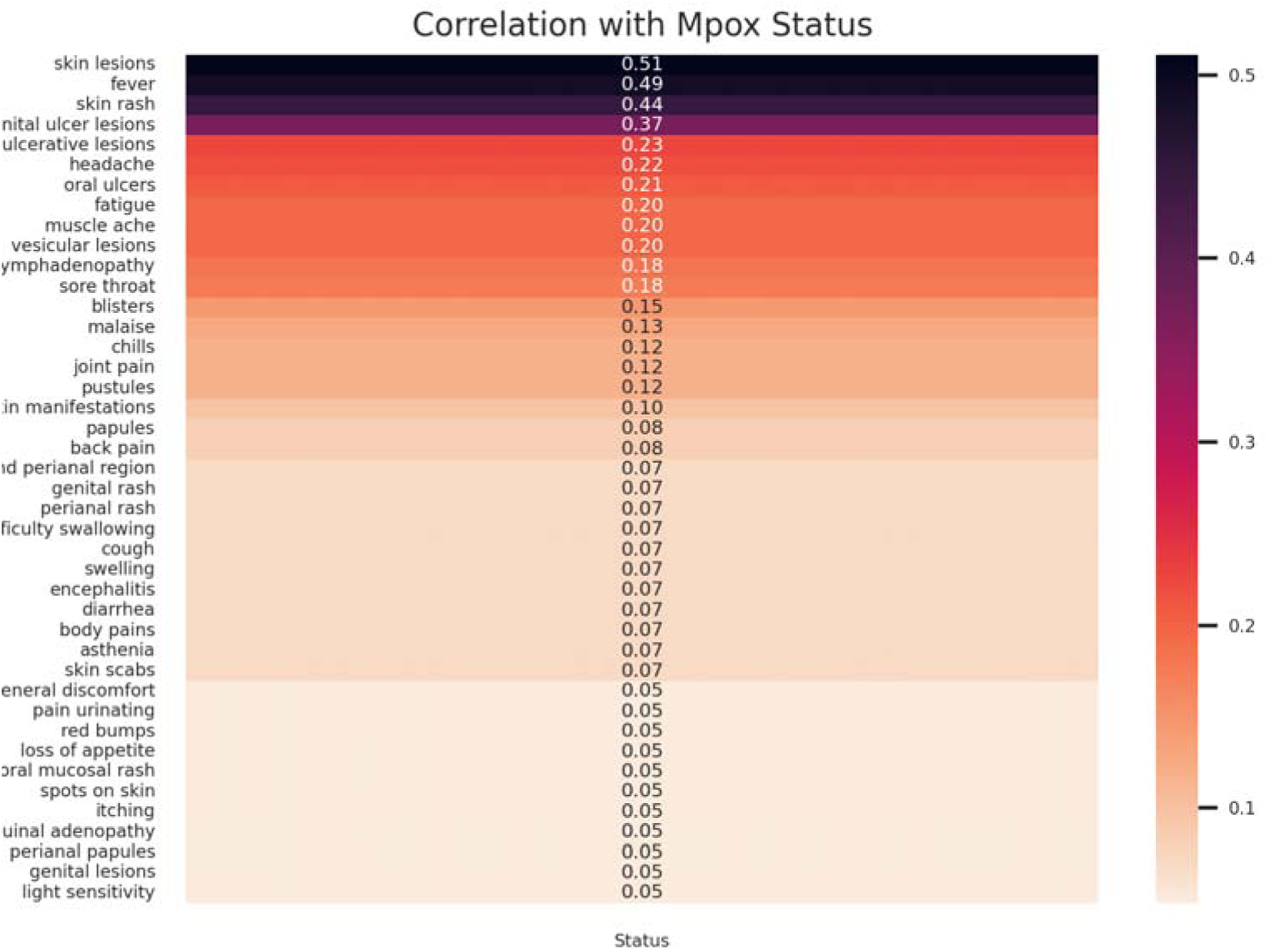
Correlation matrix of features (symptoms) with Mpox status. Skin lesions, fever and skin rash are shown to have the strongest positive correlation values, and graded in intense purple coloured rows.

### Evaluation of Machine Learning Classifiers

An initial screening of machine learning models was performed using the LazyPredict library, which benchmarks various classifiers based on multiple performance metrics including accuracy, F1-score, ROC-AUC, and computation time [23]. Models showing high predictive power were selected for further refinement.

Hyperparameter tuning for the selected models was executed using GridSearchCV with cross-validation, optimizing performance parameters and reducing the risk of overfitting [24].

## Results

### Exploratory Data Analysis

A global overview of Mpox case distribution identified the top 10 most affected countries, in descending order: the Democratic Republic of Congo (DRC), Canada, Nigeria, Portugal, Chile, Singapore, Panama, South Africa, Burundi, and Argentina. Among these, DRC and Burundi—both African nations—are currently experiencing outbreaks linked to the Clade I lineage. Other countries within the Clade Ib lineage include Uganda, Kenya, Rwanda, and Gabon (Figure 3).

**Fig 3A).**
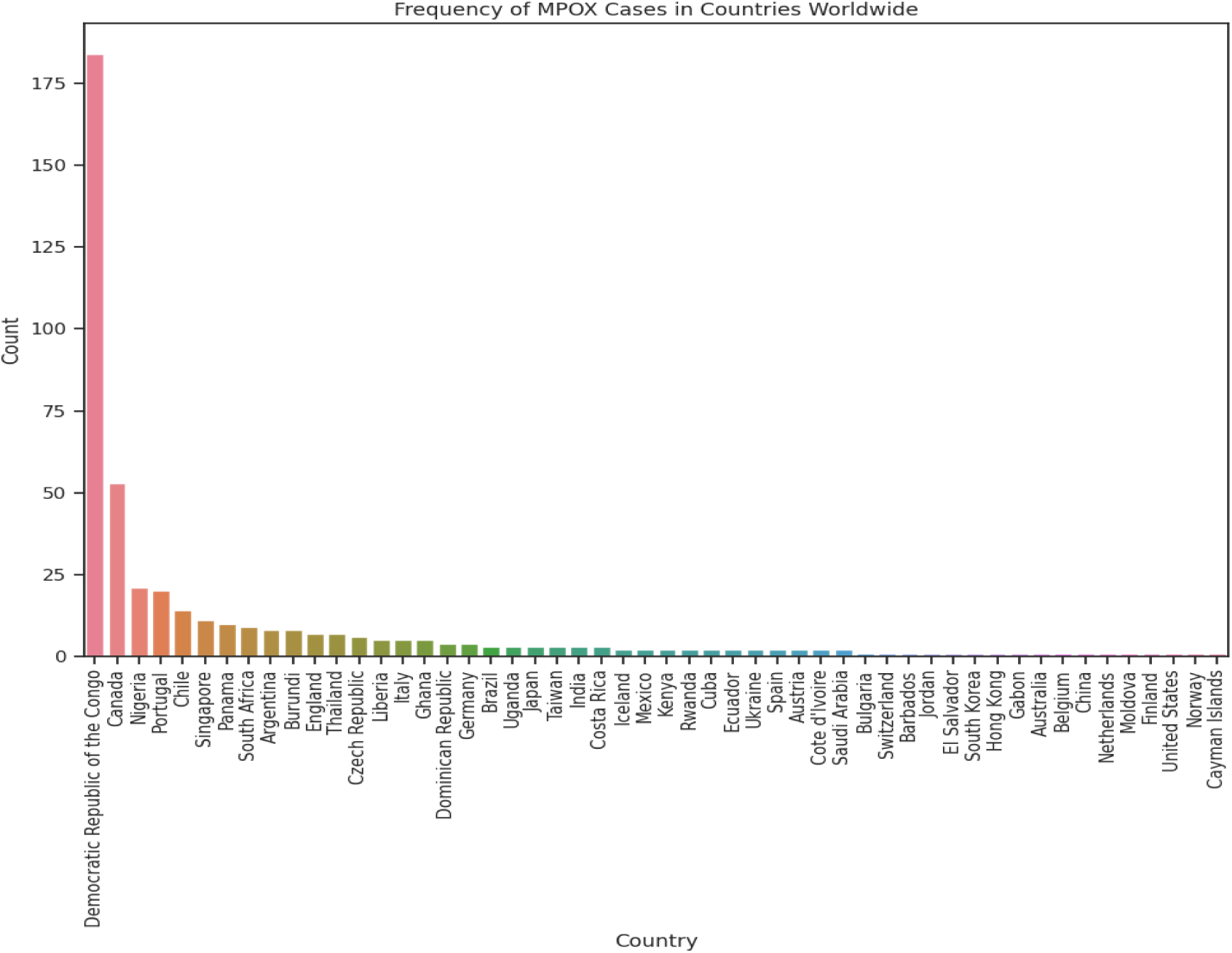
Frequency Distribution of Mpox Cases Worldwide. The DRC, Canada, Nigeria, Portugal and Chile have the highest cases in our dataset.

**Fig 3B:**
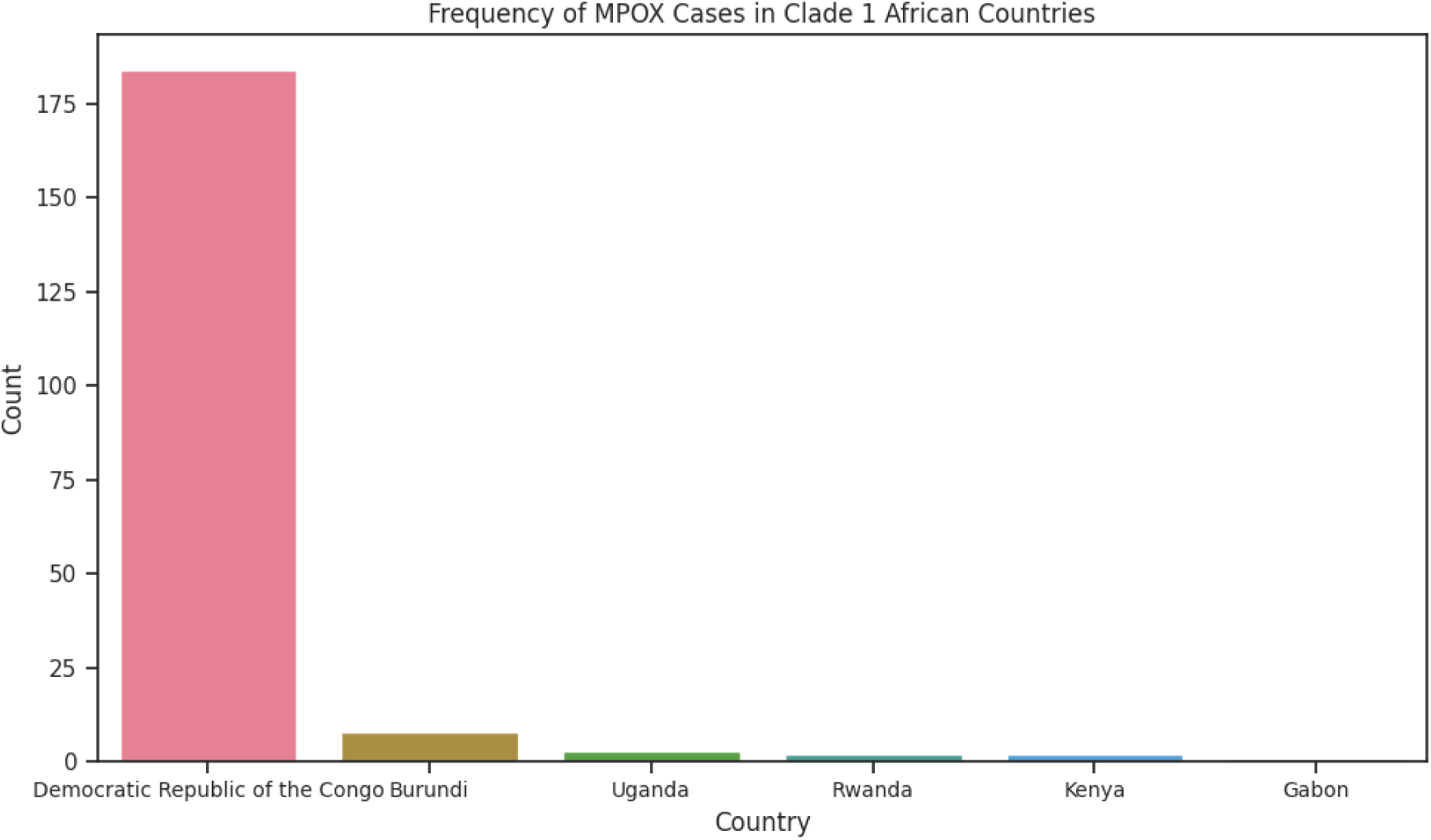
Frequency of Mpox cases in Clade 1 African Countries. The DRC records significantly highest cases among other countries in Africa represented in our dataset.

Demographic analysis revealed a higher proportion of Mpox cases among males compared to females, suggesting a possible sex-based predisposition potentially related to transmission dynamics (Figure 4B). Age stratification (Figure 4C) showed that individuals aged 20–64 years accounted for the largest share of cases, followed by the 0–19 and ≥65 age groups.

**Fig 4:**
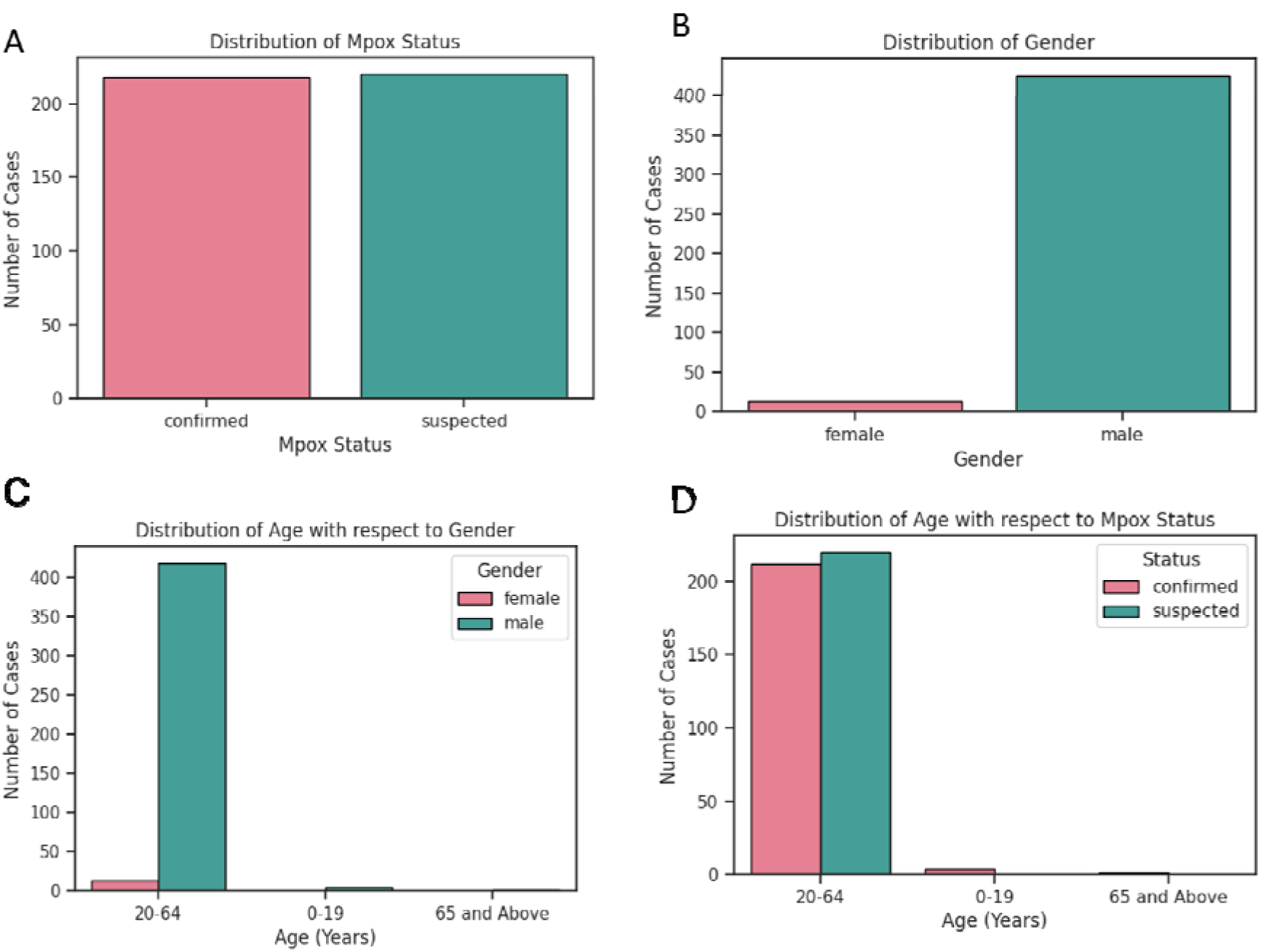
Exploratory Data Analysis. A) Distribution of Mpox Status (B) Distribution of gender in the dataset. There exists an approximately equal distribution of confirmed and suspected Mpox cases which potentially deals with class imbalance. Males show significant numbers of cases than females. (C) Distribution of Age with respect to Gender and (D) Distribution of Age with respect to Mpox status. Males between the ages 20 – 64 show the highest frequency of Mpox cases. Also, there is an almost equal distribution between confirmed and suspected Mpox cases among individuals aged 20 – 64 years.

### Correlation Analysis

Correlation analysis was performed to identify clinical features most strongly associated with Mpox status to inform model development. The highest inter-feature correlation was observed between “skin scabs” and “skin spots” (Pearson’s r = 0.71; Figure 2A). Despite this strong correlation, both features were retained in the model due to their potential to provide complementary predictive value. Regarding their association with Mpox status, “skin lesions” exhibited the strongest positive correlation (r = 0.51), followed by “fever” (r = 0.49) and “skin rash” (r = 0.44) (Figure 2B).

### Supervised Machine Learning

A total of 26 supervised machine learning algorithms were initially evaluated using the LazyPredict library (Figure 5). Based on classification accuracy and computational efficiency, five top-performing models were selected for further evaluation: Quadratic Discriminant Analysis (QDA), Extra Trees Classifier (ETC), Perceptron, Decision Tree Classifier (DTC), and Support Vector Classifier (SVC). Model performance was assessed across multiple metrics, including accuracy, F1-score, ROC-AUC, recall, and confusion matrices (Table 3, Figure 7).

**Fig 5:**
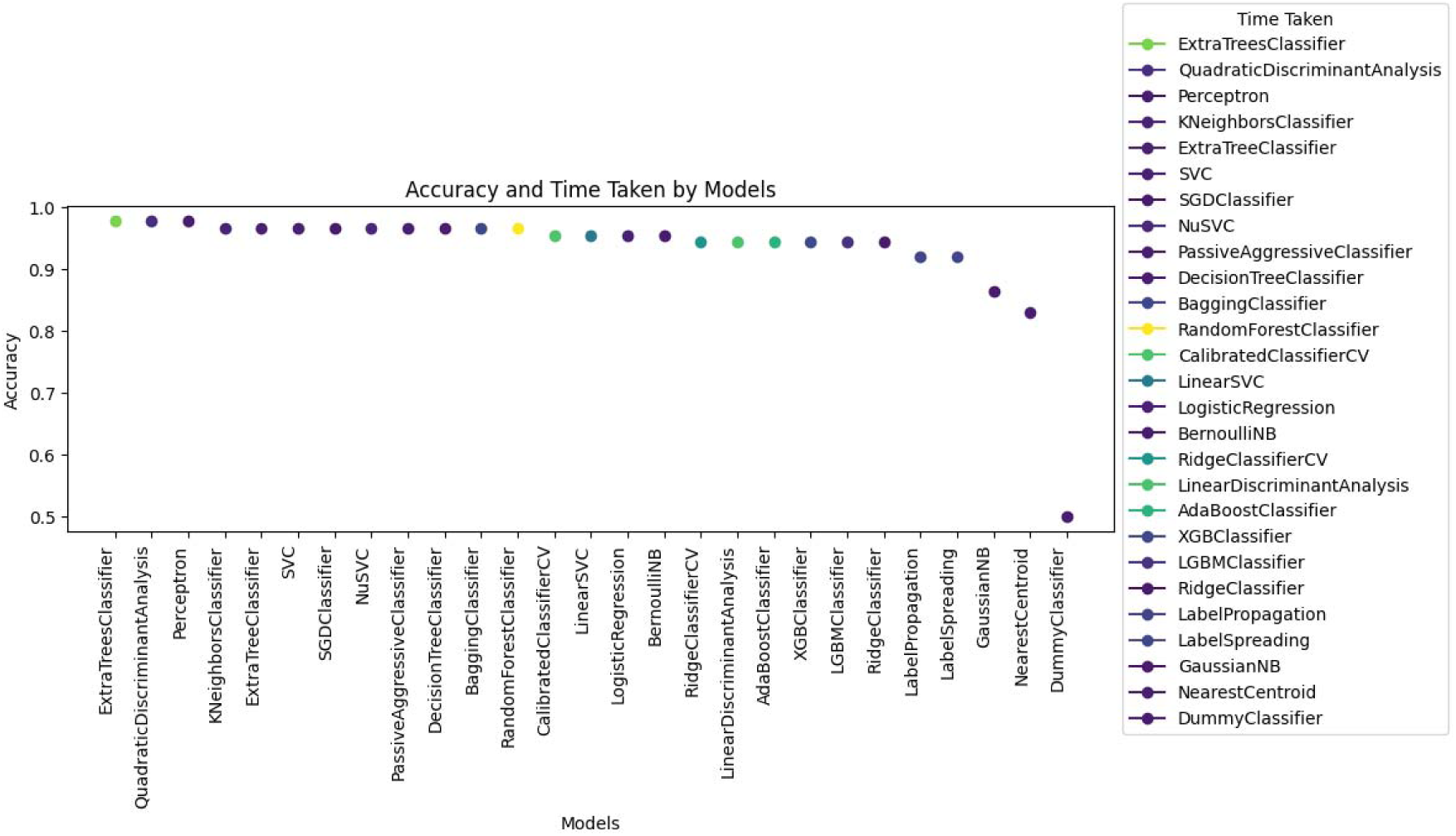
Accuracy Scores of Machine Learning Models by LazyPredict.

**Table 2.** Performance comparison of machine learning classifiers.

| Model | Accuracy | Balanced Accuracy | ROC AUC | F1 Score | Time Taken |
| --- | --- | --- | --- | --- | --- |
| ExtraTreesClassifier | 0.977272727 | 0.977272727 | 0.977272727 | 0.977260982 | 0.133304358 |
| QuadraticDiscriminantAnalysis | 0.977272727 | 0.977272727 | 0.977272727 | 0.977260982 | 0.040938377 |
| Perceptron | 0.977272727 | 0.977272727 | 0.977272727 | 0.977260982 | 0.023663044 |
| AdaBoostClassifier | 0.965909091 | 0.965909091 | 0.965909091 | 0.965869425 | 0.135079622 |
| SVC | 0.965909091 | 0.965909091 | 0.965909091 | 0.965869425 | 0.025816441 |
| SGDClassifier | 0.965909091 | 0.965909091 | 0.965909091 | 0.965869425 | 0.070972919 |
| DecisionTreeClassifier | 0.965909091 | 0.965909091 | 0.965909091 | 0.965869425 | 0.020679951 |
| RandomForestClassifier | 0.965909091 | 0.965909091 | 0.965909091 | 0.965869425 | 0.32523942 |
| ExtraTreeClassifier | 0.965909091 | 0.965909091 | 0.965909091 | 0.965869425 | 0.01622653 |
| KNeighborsClassifier | 0.965909091 | 0.965909091 | 0.965909091 | 0.965869425 | 0.021461248 |
| PassiveAggressiveClassifier | 0.965909091 | 0.965909091 | 0.965909091 | 0.965869425 | 0.026709557 |
| NuSVC | 0.965909091 | 0.965909091 | 0.965909091 | 0.965869425 | 0.052555799 |
| BaggingClassifier | 0.965909091 | 0.965909091 | 0.965909091 | 0.965869425 | 0.042726755 |
| LogisticRegression | 0.954545455 | 0.954545455 | 0.954545455 | 0.954451346 | 0.052870989 |
| LinearSVC | 0.954545455 | 0.954545455 | 0.954545455 | 0.954451346 | 0.10254693 |
| CalibratedClassifierCV | 0.954545455 | 0.954545455 | 0.954545455 | 0.954451346 | 0.194104671 |
| BernoulliNB | 0.954545455 | 0.954545455 | 0.954545455 | 0.954451346 | 0.016823292 |
| LinearDiscriminantAnalysis | 0.943181818 | 0.943181818 | 0.943181818 | 0.942997798 | 0.157581806 |
| RidgeClassifier | 0.943181818 | 0.943181818 | 0.943181818 | 0.942997798 | 0.037009954 |
| RidgeClassifierCV | 0.943181818 | 0.943181818 | 0.943181818 | 0.942997798 | 0.339598417 |
| XGBClassifier | 0.943181818 | 0.943181818 | 0.943181818 | 0.942997798 | 0.101504087 |
| LGBMClassifier | 0.943181818 | 0.943181818 | 0.943181818 | 0.942997798 | 0.091071844 |
| LabelSpreading | 0.931818182 | 0.931818182 | 0.931818182 | 0.931499741 | 0.033649921 |
| LabelPropagation | 0.931818182 | 0.931818182 | 0.931818182 | 0.931499741 | 0.027289629 |
| GaussianNB | 0.863636364 | 0.863636364 | 0.863636364 | 0.861052632 | 0.01837492 |
| NearestCentroid | 0.829545455 | 0.829545455 | 0.829545455 | 0.82444474 | 0.062399387 |
| DummyClassifier | 0.5 | 0.5 | 0.5 | 0.333333333 | 0.015606165 |

**Table 3:** Evaluation Metrics in Comparative Performance of Machine Learning Models.

| **Model** | Accuracy | F1-score | Recall | ROC-AUC |
| --- | --- | --- | --- | --- |
| **Extra Trees Classifier** | 95.5 | 95.2 | 90.9 | 95.5 |
| **Perceptron** | 97.7 | 97.7 | 95.5 | 97.7 |
| **Quadratic Discriminant Analysis** | 97.7 | 97.7 | 95.5 | 97.7 |
| **Decision Tree Classifier** | 94.3 | 93.9 | 88.6 | 94.3 |
| **Support Vector Classifier** | 97.7 | 97.7 | 95.5 | 97.7 |

After hyperparameter tuning with GridSearchCV, the ETC model achieved an accuracy and ROC-AUC of 95.5%, with an F1-score of 95.2%. The DTC model recorded an accuracy of 94.3%, ROC-AUC of 94.3%, and an F1-score of 93.9%. Notably, the Perceptron, QDA, and SVC models each attained superior results, with accuracy, F1-score, and ROC-AUC all reaching 97.7%. Regarding recall, ETC and DTC achieved 90.9% and 88.6%, respectively, while SVC, Perceptron, and QDA outperformed them with a recall of 95.5%. Confusion matrix analyses provided detailed insights into each model’s ability to differentiate between suspected (label = 0) and confirmed (label = 1) Mpox cases.

### Feature Importance

Permutation importance analysis was conducted using the best-performing models, SVC and QDA, to determine the most influential symptoms for Mpox classification (Figure 6). “Skin rash” emerged as the most impactful feature, with a permutation importance (PI) score of approximately 0.12. “Skin lesions” and “fever” followed closely, each with a PI score of 0.11. These results underscore the critical role of dermatological symptoms in the diagnosis and classification of Mpox.

**Fig 6:**
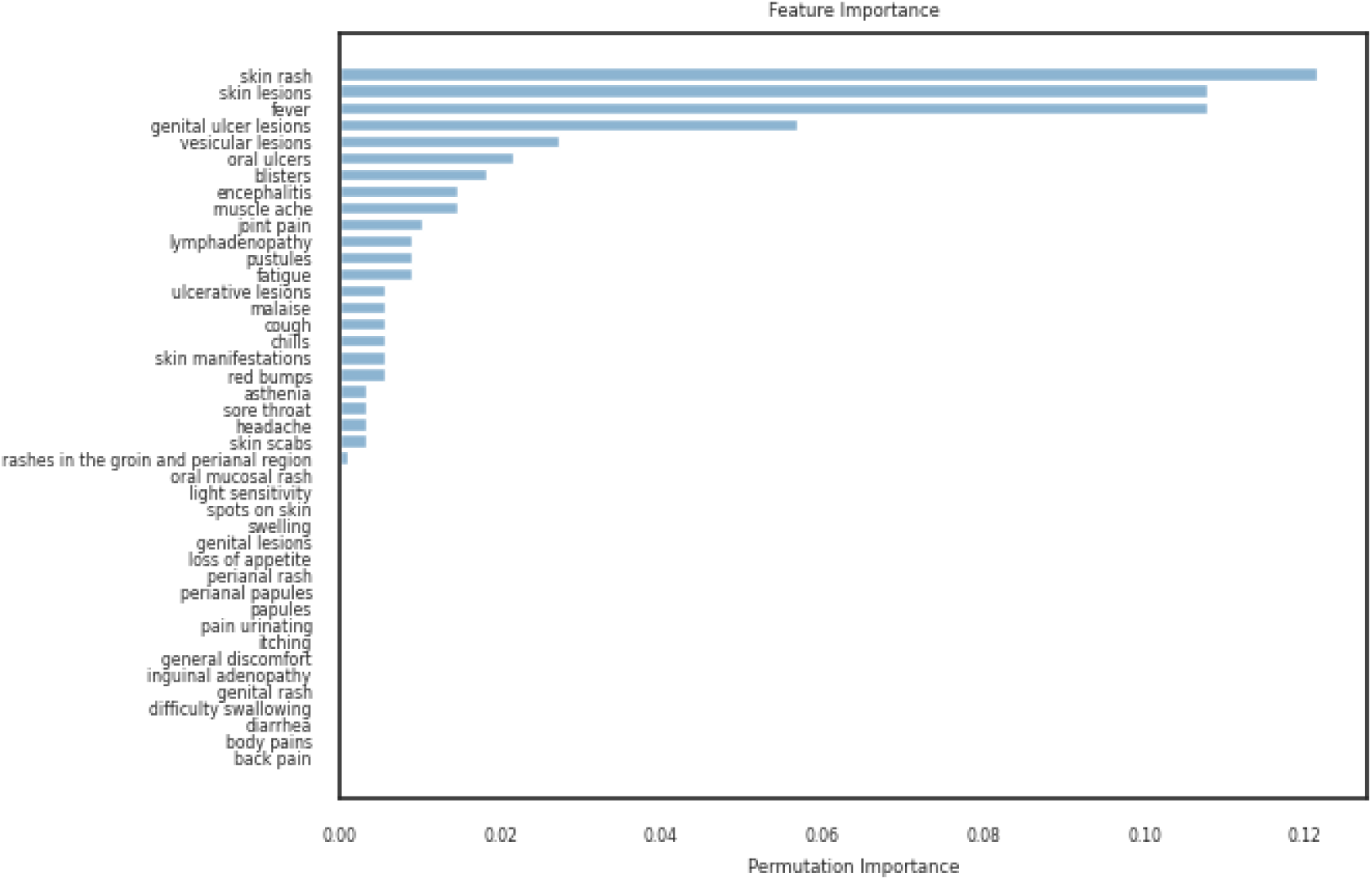
Feature Importance Analysis of Mpox Symptoms. Skin rash gives the highest PI score of 0.12, followed by skin lesions (0.11) and fever (0.11), highlighting their influence on the models’ predictions.

**Fig 7:**
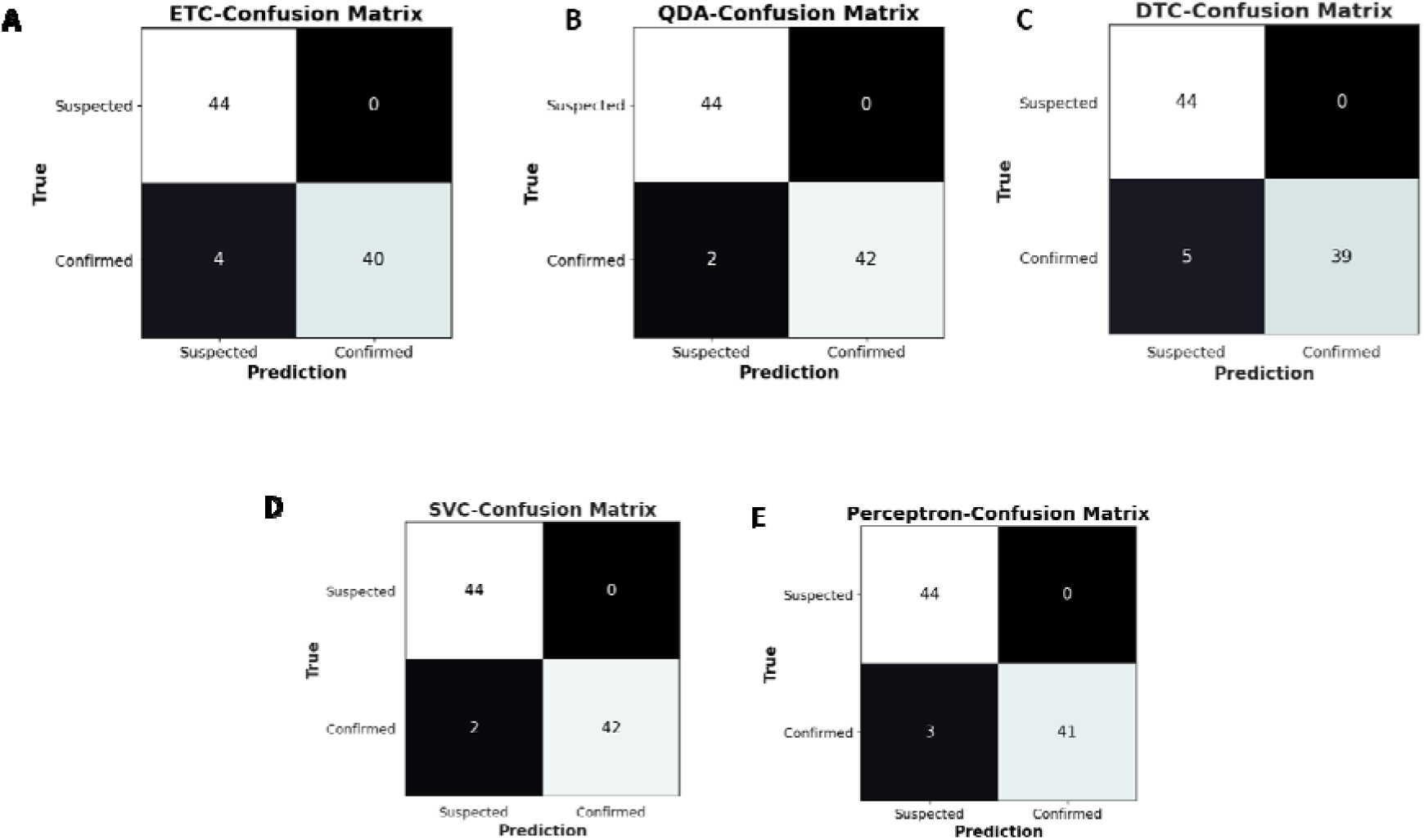
Confusion matrices of Machine Learning models. **A)** ExtraTrees Classifier (B) Quadratic Discriminant Analysis (C) Decision Tree Classifier (D) Support Vector Classifier (E) Perceptron

## Discussion

The global transmission dynamics of Mpox have shifted markedly in recent years, driven by increased international travel, urbanization, and global interconnectedness, which have facilitated viral spread to non-endemic regions [2, 25]. In Africa, the resurgence of Mpox can be attributed to waning population immunity due to the cessation of smallpox vaccination programs, fragile healthcare infrastructure, and protracted conflict disrupting healthcare access [26].

A major barrier to effective Mpox control remains underreporting. Factors contributing to this include the limited availability of molecular diagnostics, shortages in test kits, and persistent disruptions to routine healthcare delivery, especially in rural and conflict-prone areas [27]. In addition, asymptomatic and subclinical infections—estimated to occur in 1.8–6.5% of individuals—may further obscure the true burden of Mpox [28]. Inconsistent documentation of symptoms by healthcare providers and variability in clinical presentation highlight the pressing need for improved frontline training and standardized case reporting protocols [29, 30].

The recent emergence and spread of Clade 1b—predominantly associated with sexual transmission—have led to outbreaks in regions bordering the Democratic Republic of Congo (DRC), including Kenya, Burundi, Rwanda, and Uganda, which had previously reported few or no cases [31, 32]. This is concerning given the higher transmissibility and virulence associated with Clade I strains [8, 33]. Children in rural African communities have historically experienced the greatest Mpox-related morbidity and mortality [34], a trend supported by 2023 surveillance data from the DRC and Burundi, where individuals under 15 years represented a substantial proportion of cases [35].

Interestingly, our study found that adults aged 20–64 years accounted for the highest number of cases, which contrasts with prior African surveillance data. This discrepancy may stem from demographic data imputation methods or reflect genuine age-related variation in transmission dynamics across regions and outbreak contexts. In high-income, non-endemic countries like Canada, Portugal, and Chile, prior outbreak data have similarly shown Mpox to be most prevalent among adults in this age range [36, 37]. It is also possible that reporting biases exist, with more severe pediatric cases more likely to be identified and recorded in endemic settings, while milder or self-limiting adult cases may dominate in high-surveillance environments [38].

Sex-based disparities were also evident. Our findings indicate a higher incidence among males, which aligns with WHO reports from the 2022 and 2023 outbreaks that identified men—particularly men who have sex with men (MSM)—as a high-risk group [4, 39]. Transmission through sexual networks has been a significant driver of recent outbreaks, and MSM populations have been shown to be disproportionately affected [40–42]. Behavioral, social, and structural factors likely contribute to this risk stratification and underscore the importance of targeted public health messaging.

To optimize machine learning performance, we performed a correlation analysis of clinical features, identifying “skin lesions” and “skin scabs” as the most strongly correlated symptoms. Despite their collinearity (r = 0.71), both features were retained due to their potential additive value in classification performance. Feature retention decisions must balance overfitting risk against the loss of clinically relevant information, particularly in noisy real-world datasets [43].

From an algorithmic standpoint, the Quadratic Discriminant Analysis, Support Vector Classifier, and Perceptron models achieved the highest performance across multiple metrics, including accuracy (97.7%), F1-score, recall, and ROC-AUC. While accuracy is a commonly cited metric, it alone is insufficient to evaluate a model’s real-world utility—especially in imbalanced or clinical datasets. Therefore, we also relied on confusion matrices, which provide a more detailed view of model behavior by quantifying true/false positives and negatives [44, 45].

Our confusion matrix analysis revealed that all models correctly classified 44 true positives with zero false positives; however, SVC and QDA yielded the fewest false negatives (n = 2), reflecting superior sensitivity. High recall is critical in infectious disease diagnostics, as it ensures that true cases are not overlooked—a key priority during public health outbreaks. These findings support the use of QDA and SVC as clinically reliable diagnostic models.

Importantly, both QDA and SVC have been successfully applied in various medical contexts. QDA has been utilized in the classification of coronary heart disease [46] and keratoconus [47], while SVC has demonstrated utility in diagnosing diabetes [48], multiple cancers [49, 50], heart disease [51], and COVID-19 [52]. Its adaptability for both clinical and imaging applications also makes SVC particularly attractive in digital health settings [53, 54].

To interpret model decision-making, we performed permutation-based feature importance analysis. “Skin rash,” “skin lesions,” and “fever” emerged as the top three features across models, this is unsurprising given their central role in Mpox presentation. However, “lymphadenopathy,” a hallmark feature distinguishing Mpox from other vesiculopustular illnesses (e.g., smallpox, varicella), ranked only 11th, with a permutation score below 0.02. This unexpected result suggests that lymphadenopathy, while clinically significant, may not be consistently reported or may lack sufficient discriminatory power in our dataset. This has implications for the weighting of clinical features in future algorithm design and suggests that data quality plays a central role in driving model interpretability and accuracy [55].

Despite these promising results, our study has several limitations. The presence of missing demographic data necessitated statistical imputation, which may have introduced biases, especially in age- and gender-stratified analyses. Furthermore, the dataset may not fully represent the global Mpox burden due to uneven reporting practices and underdiagnosis in low-resource settings. The retrospective, observational nature of our analysis precludes causal inference. Furthermore, symptom data were largely self-reported or extracted from heterogeneous health systems, introducing potential variability and recall bias.

Future work should prioritize the prospective collection of standardized, high-quality clinical data from both endemic and non-endemic regions. Integrating behavioral risk factors, laboratory confirmations, and virological profiles could further enhance the predictive power of ML models. Real-world implementation studies are also needed to evaluate the usability and diagnostic usefulness of SVC and QDA models, particularly in settings with limited infrastructure. Finally, combining symptom-based models with genomic or image-based diagnostic tools may improve classification in complex clinical scenarios, aiding in early detection and outbreak containment.

## Conclusions

This study demonstrated the potential of machine learning models for classifying Mpox cases based on clinical symptoms, with Quadratic Discriminant Analysis (QDA), Support Vector Classification (SVC), and Perceptron emerging as the most effective classifiers. Our analysis also highlighted important epidemiological trends, including the predominance of Mpox in males and individuals aged 20–64 years, which demonstrated notable geographic disparities in case distribution. Although the models performed well, larger and more standardized datasets are needed for validation. Future research should incorporate temporal, geospatial, genomic, and behavioral data for more robust predictive tools.

## Data Availability

The datasets supporting the analysis in this article are openly accessible and available at https://github.com/globaldothealth/outbreak-data/wiki/GHL2024.D11.1E71 and https://github.com/globaldothealth/monkeypox

https://github.com/globaldothealth/monkeypox

## List of Abbreviations

AI: Artificial Intelligence
DRC: Democratic Republic of Congo
DTC: Decision Trees Classifier
ETC: ExtraTrees Classifier
ML: Machine Learning
MPXV: Monkeypox Virus
MSM: Men who have sex with men
PCR: Polymerase Chain Reaction
QDA: Quadratic Discriminant Analysis
ROC-AUC: Receiver Operating Characteristics- Area under the Curve
SVC: Support Vector Classifier
SVM: Support Vector Machine
WHO: World Health Organization

## Declarations

### Clinical trial number

Not Applicable

## Ethics approval and consent to participate

Not Applicable

## Consent for publication

Not Applicable

## Authors’ contributions

SCO and KO conceived and designed the study, SCO implemented data analysis, SCO and KO drafted the manuscript, SCO, MO and FL edited the manuscript. All authors read, revised and approved the final manuscript.

## Acknowledgements

We are thankful to the students and staff of the Centre for Genomics Research in Biomedicine, Mountain Top University for the clerical support.

